# Association of healthcare fragmentation and overall survival in patients with kidney transplant in Colombia

**DOI:** 10.1101/2024.12.12.24318934

**Authors:** Luis Manuel Barrera, Daniela Sánchez-Santiesteban, Giancarlo Buitrago

## Abstract

**Objective:** Kidney transplantation requires a multidisciplinary approach to achieve optimal outcomes. Healthcare fragmentation can negatively impact clinical outcomes; however, this issue remains understudied in low- and middle-income countries (LMICs). This study aimed to assess healthcare fragmentation in kidney transplant patients during their first post-transplant year and evaluate its association with three-year survival among patients enrolled in Colombia’s contributory healthcare scheme.

**Methods:** A retrospective cohort study was conducted using administrative data from Colombia’s contributory healthcare scheme. The cohort included kidney transplant recipients (2012–2016) who survived the first post-transplant year. Healthcare fragmentation was measured by the number of unique providers involved in the first year. Patients were categorised into high- and low-fragmentation groups based on the 75th percentile of provider distribution. The primary outcome was three-year survival, analysed using multivariate Cox regression to estimate hazard ratios (HRs), adjusted for age, sex, Charlson Comorbidity Index (CCI), insurer, region, and transplant year.

**Results:** The cohort comprised 2,028 kidney transplant patients, with a mean age of 47.7 years (SD: 13.4), 38.7% female, and 68.7% presenting a CCI≤3. Healthcare fragmentation ranged from 1 to 34 providers, with a mean of 8.94 (SD: 6.77). High fragmentation (≥11 providers) was observed in 30.2% of patients. Three-year mortality was significantly higher in the high-fragmentation group (18%) compared to the low-fragmentation group (12%) (p=0.04). High fragmentation was associated with a 49% increased mortality risk (adjusted HR: 1.49; 95% CI: 1.12–1.97; p=0.01).

**Conclusion:** Healthcare fragmentation significantly reduces three-year overall survival in kidney transplant recipients in Colombia. These findings underscore the importance of integrated care models and improved coordination among providers to enhance patient outcomes, particularly in resource-limited settings.

## INTRODUCTION

Kidney transplantation is recognised as the most effective treatment for patients with end-stage renal disease, significantly improving survival rates and quality of life compared to long-term dialysis [1,2]. Despite its clinical benefits, the success of renal transplantation depends heavily on comprehensive, multidisciplinary care that addresses various challenges, including immunosuppressive therapy, management of comorbid conditions, and the prevention and early detection of complications [3–5]. Coordinated care is critical in ensuring optimal outcomes, as patients require regular follow-ups, monitoring by specialised teams, and laboratory by healthcare providers [5]. However, fragmentation within healthcare systems, defined by inadequate communication and lack of integration among providers, disrupts this continuum of care, leading to adverse clinical outcomes [6–9]. In kidney transplantation, for example, fragmentation can lead to delays in diagnosis and treatment, duplication of medical services, increased costs, and worse health outcomes, ultimately compromising the potential benefits of transplantation [10,11].

Due to their fragmented health systems, Latin American countries face significant challenges in delivering integrated healthcare. In Colombia, fragmentation manifests both horizontally, through a lack of coordination between providers at the same level of care, and vertically [12]. This fragmentation is further exacerbated in Colombia by the decentralised structure of the healthcare system, which disperses responsibility across multiple actors, including public and private providers, insurers, and regional authorities. For complex medical procedures like kidney transplantation, the consequences of fragmentation are particularly severe as they encounter obstacles such as inconsistent follow-up, limited access to necessary medications, and insufficient communication between transplant centres and other providers. These challenges not only hinder recovery but also increase the likelihood of complications, hospital readmissions, and preventable mortality.

Healthcare fragmentation has been studied in chronic conditions such as cancer and cardiovascular diseases [13–17]; however, kidney transplantation, which requires a similarly high level of care integration, remains understudied. To our knowledge, no studies have specifically evaluated the association between healthcare fragmentation and survival outcomes in renal transplant recipients in low- and middle-income countries (LMICs). This gap in the literature underscores the need for targeted research to understand better how fragmented care influences transplant success, particularly in the Colombian context, where fragmentation challenges are pronounced.

This study aims to address this gap by assessing the extent of care fragmentation experienced by renal transplant patients during the critical first year following transplantation and evaluating its association with three-year overall survival. By analysing data from a large national cohort, this research seeks to provide evidence of the association of healthcare fragmentation on clinical outcomes. The findings can potentially inform health policies and interventions to reduce care fragmentation, improve coordination among healthcare providers, and optimise resource use in this population. Ultimately, the study highlights the importance of integrated care models in enhancing renal transplant patients’ survival and quality of life, particularly in resource-limited and fragmented healthcare systems like Colombia.

## METHODS

### Study Design and Population

This retrospective cohort study was conducted based on administrative data of adult patients who underwent kidney transplantation between 2012 and 2016 and were enrolled in the contributory health insurance system. The cohort was constructed using the Base for the Study of Capitation Unit Sufficiency (UPC) to identify all adult patients with a kidney transplantation record [18]. All subjects included in the study had to survive the first-year post-transplant to allow for the measurement of fragmentation during this period. After this initial year, each subject was followed for up to three years or until death, whichever occurred first. We used the Mortality Registry Module from the Unified Affiliation Registry (RUAF) to determine the mortality date [19].

This study utilised two primary administrative databases, anonymised and linked, using unique identifiers by the Ministry of Health (MoH). The UPC database is highly standardised and is the primary source of information used by the Ministry of Health (MoH) for the annual estimation of the UPC in the healthcare system. The database includes detailed information on each utilised service, such as the Unified Health Procedure Code (CUPS), an ICD-10 code associated with the service, the service date, the cost paid by each insurer to each provider, the patient’s sex, the insurer to which the patient is affiliated, the city where the service was provided, the provider’s registration code, and an anonymised individual identifier for each affiliate. Additionally, the RUAF Mortality Registry Module contains information from death certificates for all Colombians, including the date of death and diagnosis. International assessments have confirmed the reliability of RUAF data, with 91% of deaths registered through death certificates as of 2016 [20]. These databases were provided to the Clinical Research Institute at the Faculty of Medicine of the Universidad Nacional de Colombia by the Office of Information Technologies of MoH for research purposes. This study was approved by the Faculty of Medicine Ethics Committee at the Universidad Nacional de Colombia (Approval Number: 004-029).

### Exposure, Outcome, and Control Variables

As mentioned, all included patients survived the first-year post-transplantation as an inclusion criterion. To measure healthcare fragmentation, we counted the number of different healthcare providers involved in the patient’s care during the first year of post-transplantation. This number was the primary exposure variable and has been used in similar studies [13–16,21]. After the first year of survival, all patients were followed for an additional three years. Using information from the RUAF database, the three-year survival time was estimated, corresponding to the study’s primary outcome variable. Control variables included age, sex, comorbidities measured using the Charlson Comorbidity Index (CCI) [22], insurer, geographic region, and year of transplantation.

### Analysis

Baseline characteristics of the cohort were described using measures of central tendency and dispersion for continuous variables and relative and absolute frequencies for categorical variables. The quartiles of the primary exposure variable (i.e., the number of different providers in the first-year post-transplantation) were identified.

The cohort was divided into high fragmentation (those exposed to a number of providers greater than or equal to the 75th percentile of the distribution) and low fragmentation (those exposed to a number of providers below the 75th percentile). An analysis of the crude three-year mortality proportion and unadjusted survival was performed. Using a multivariate Cox regression model, the hazard ratio (HR) of being exposed to highly fragmented healthcare compared to low-fragmentation healthcare was estimated, controlling for the previously mentioned covariables. The proportionality assumption was evaluated with graphical and statistical tests using Schoenfeld residuals. All estimators were presented with 95% confidence intervals. Analyses were conducted using Stata 17 MP (licensed to the Universidad Nacional de Colombia). This article followed the STROBE (Strengthening the Reporting of Observational Studies in Epidemiology) guidelines to ensure transparency and thoroughness in reporting the findings.

## RESULTS

Between 2012 and 2016, a total of 3,999 patients underwent kidney transplantation. After excluding 47 patients under 18 years old and 1,924 patients who died within the first year, 2,028 patients were eligible for the study. No records were lost in the follow-up as each of the 2,028 participants was followed for three years or until death, whichever occurred first. The flowchart in Figure 1 provides additional details on the selection process.

**Fig 1.**
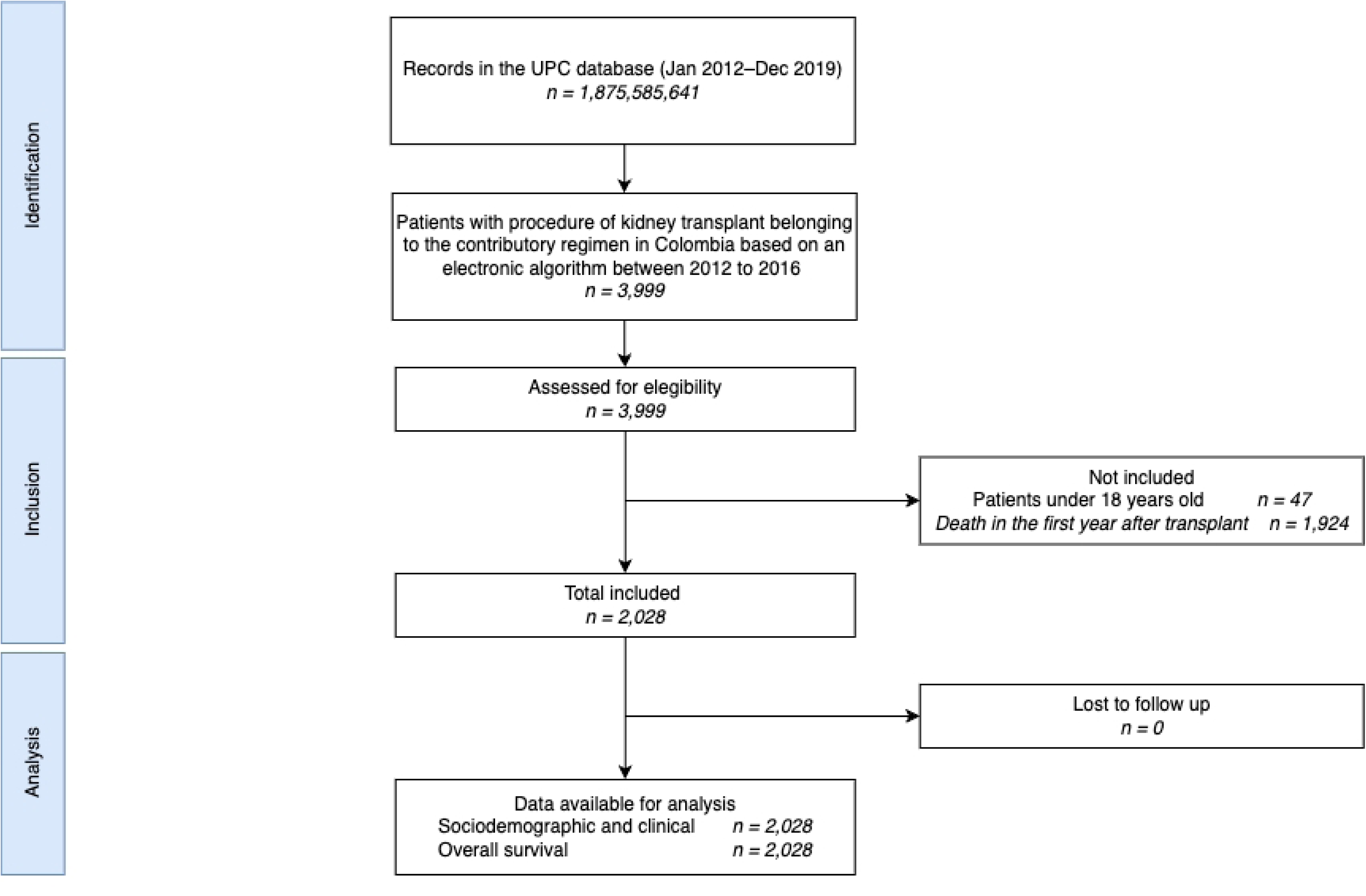
STROBE flow chart. UPC: Base for the Study of Capitation Unit Sufficiency Database.

Table 1 presents detailed sociodemographic, clinical and exposure characteristics of the cohort. Of the total cohort, 785 patients (38.7%) were women, and the mean age was 47.7 years (Standard Deviation [SD] 13.4 years). A total of 1,394 patients (68.7%) had a CCI of 3 or less. Bogotá was the region with the highest number of transplants, with a total of 923 (45.5%), and a decline in the number of transplants per year was observed (see Table 1).

**Table 1.**
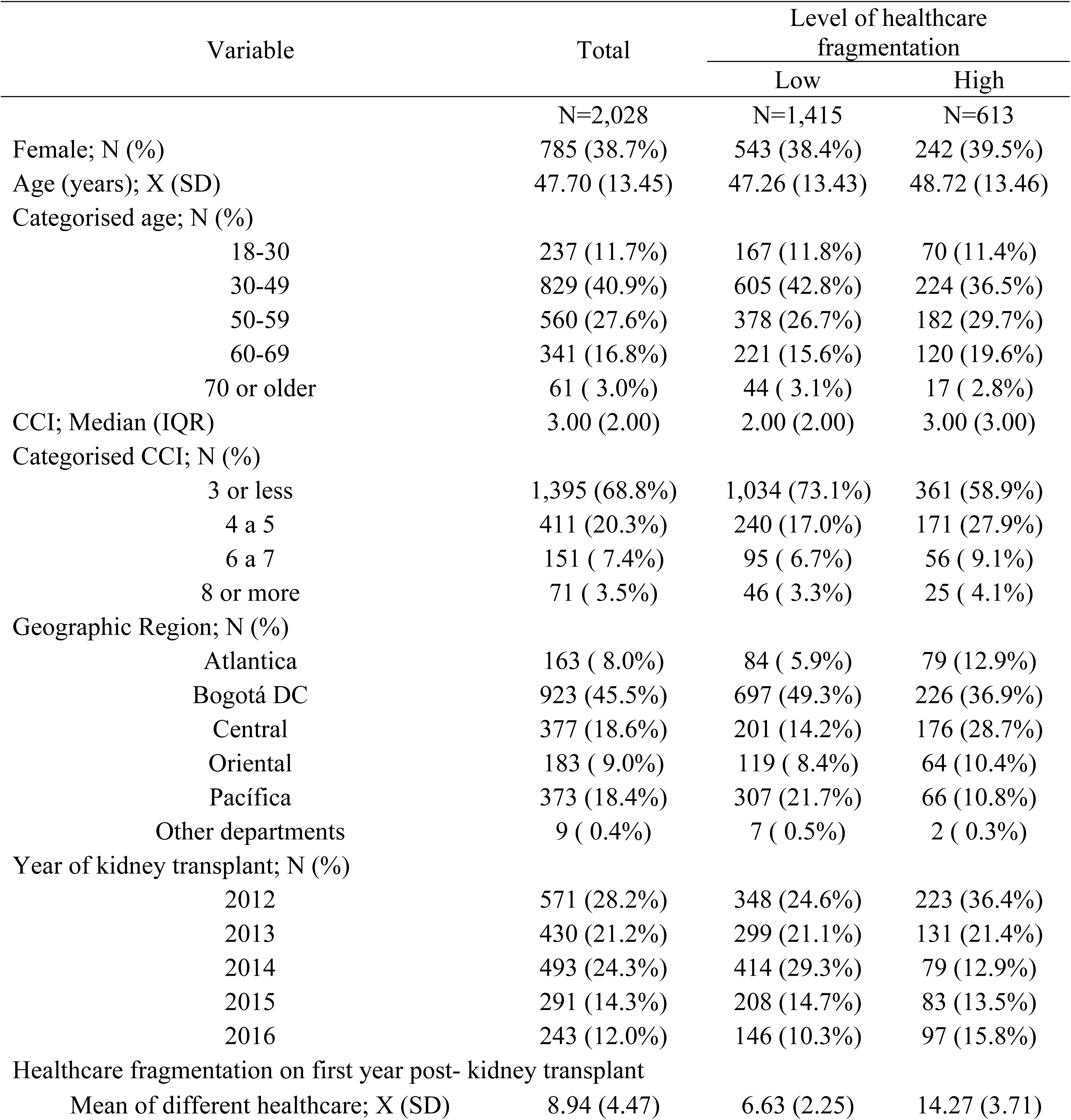
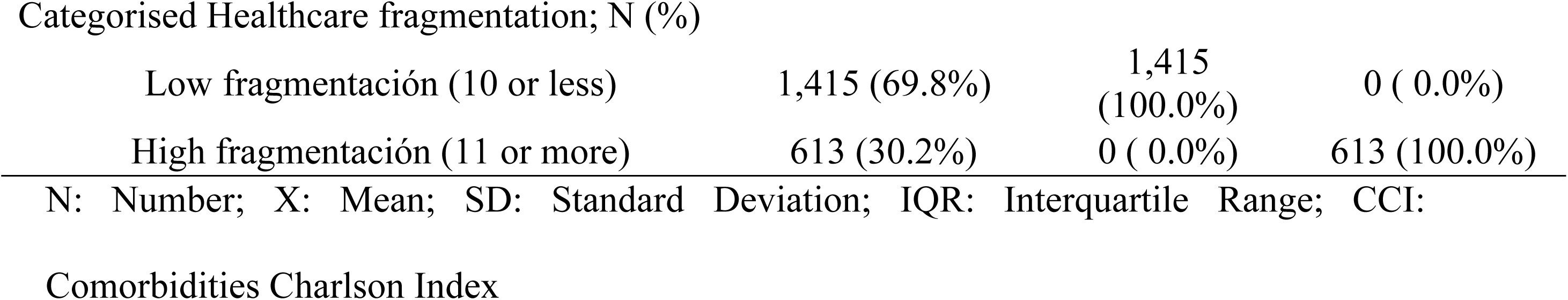
Description of cohort baseline characteristics.

| Variable | Total | Level of healthcare fragmentation |  |
| --- | --- | --- | --- |
|  |  | Low | High |
|  | N=2,028 | N=1,415 | N=613 |
| Female; N (%) | 785 (38.7%) | 543 (38.4%) | 242 (39.5%) |
| Age (years); X (SD) | 47.70 (13.45) | 47.26 (13.43) | 48.72 (13.46) |
| Categorised age; N (%) |  |  |  |
| 18-30 | 237 (11.7%) | 167 (11.8%) | 70 (11.4%) |
| 30-49 | 829 (40.9%) | 605 (42.8%) | 224 (36.5%) |
| 50-59 | 560 (27.6%) | 378 (26.7%) | 182 (29.7%) |
| 60-69 | 341 (16.8%) | 221 (15.6%) | 120 (19.6%) |
| 70 or older | 61 ( 3.0%) | 44 ( 3.1%) | 17 ( 2.8%) |
| CCI; Median (IQR) | 3.00 (2.00) | 2.00 (2.00) | 3.00 (3.00) |
| Categorised CCI; N (%) |  |  |  |
| 3 or less | 1,395 (68.8%) | 1,034 (73.1%) | 361 (58.9%) |
| 4 a 5 | 411 (20.3%) | 240 (17.0%) | 171 (27.9%) |
| 6 a 7 | 151 ( 7.4%) | 95 ( 6.7%) | 56 ( 9.1%) |
| 8 or more | 71 ( 3.5%) | 46 ( 3.3%) | 25 ( 4.1%) |
| Geographic Region; N (%) |  |  |  |
| Atlantica | 163 ( 8.0%) | 84 ( 5.9%) | 79 (12.9%) |
| Bogotá DC | 923 (45.5%) | 697 (49.3%) | 226 (36.9%) |
| Central | 377 (18.6%) | 201 (14.2%) | 176 (28.7%) |
| Oriental | 183 ( 9.0%) | 119 ( 8.4%) | 64 (10.4%) |
| Pacífica | 373 (18.4%) | 307 (21.7%) | 66 (10.8%) |
| Other departments | 9 ( 0.4%) | 7 ( 0.5%) | 2 ( 0.3%) |
| Year of kidney transplant; N (%) |  |  |  |
| 2012 | 571 (28.2%) | 348 (24.6%) | 223 (36.4%) |
| 2013 | 430 (21.2%) | 299 (21.1%) | 131 (21.4%) |
| 2014 | 493 (24.3%) | 414 (29.3%) | 79 (12.9%) |
| 2015 | 291 (14.3%) | 208 (14.7%) | 83 (13.5%) |
| 2016 | 243 (12.0%) | 146 (10.3%) | 97 (15.8%) |
| Healthcare fragmentation on first year post- kidney transplant |  |  |  |
| Mean of different healthcare; X (SD) | 8.94 (4.47) | 6.63 (2.25) | 14.27 (3.71) |
| Low fragmentación (10 or less) | 1,415 (69.8%) | 1,415<br>(100.0%) | 0 ( 0.0%) |
| High fragmentación (11 or more) | 613 (30.2%) | 0 ( 0.0%) | 613 (100.0%) |

N: Number; X: Mean; SD: Standard Deviation; IQR: Interquartile Range; CCI:
Comorbidities Charlson Index

The degree of healthcare fragmentation, measured as the number of distinct providers involved in a patient’s care during the first year post-transplant, ranged from 1 to 34, with a mean of 8.94 (SD: 6.77). The 25th, 50th, and 75th percentiles were 6, 8, and 11 providers, respectively. Among the regions analysed, the Central region exhibited the highest mean fragmentation, with an average of 10.78 providers (SD: 4.65), while the Pacific region showed the lowest, with an average of 7.96 providers (SD: 3.73) (Table 2). At the departmental level, the highest fragmentation was observed in Sucre (14.00; SD: 5.10), La Guajira (12.13; SD: 5.67), and Caquetá (13.60; SD: 5.19). Conversely, the lowest fragmentation was found in Nariño (6.33; SD: 2.65) and Arauca (6.50; SD: 2.12). Figure 2 illustrates the geographic distribution of healthcare fragmentation across Colombia, highlighting significant regional variability.

**Fig 2.**
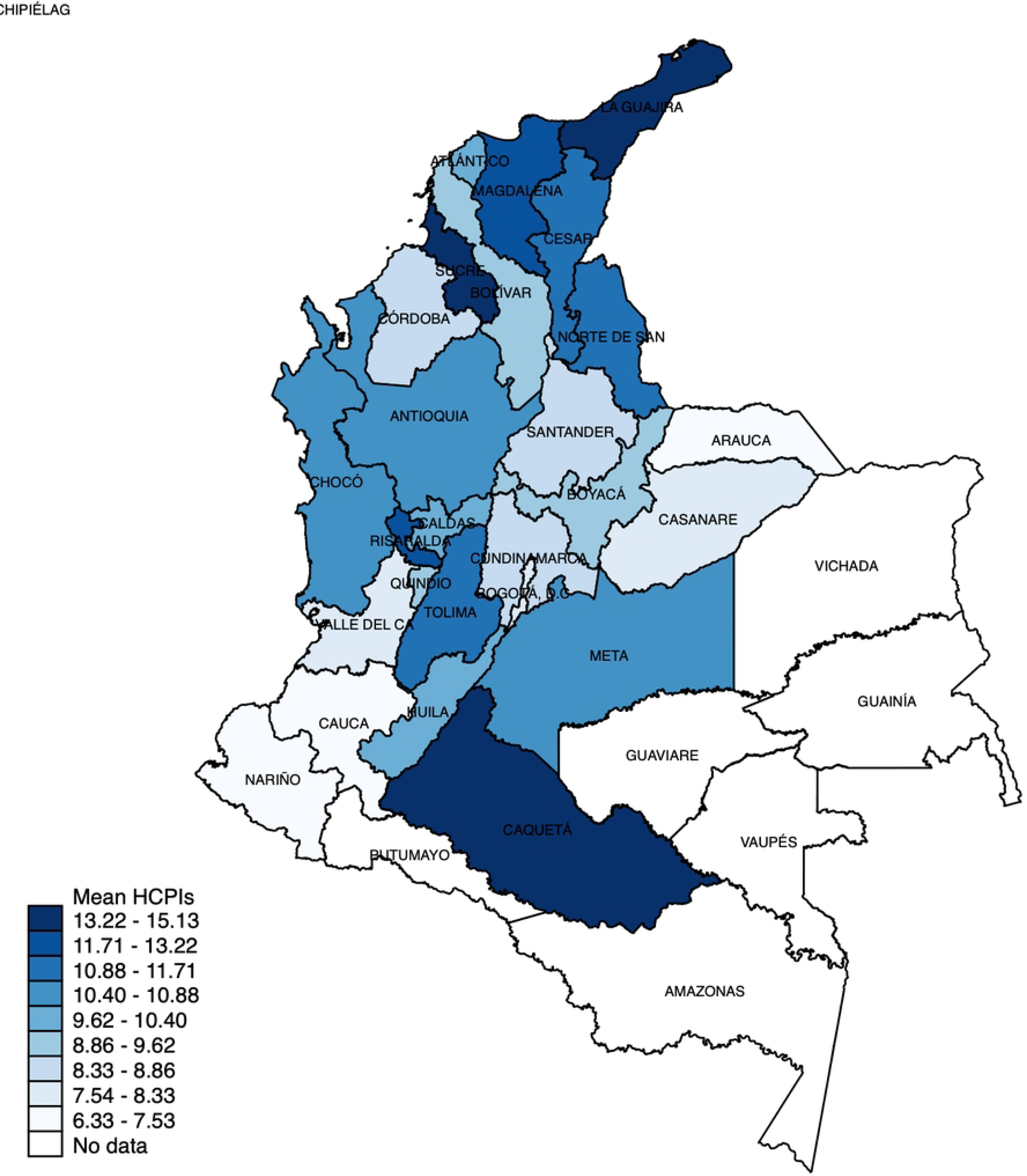
Geographical distribution of healthcare fragmentation. Geographic distribution of exposure to healthcare fragmentation measured as number of different HCPIs in the first year of care after kidney transplantation in adults belonging to the contributory regimen in Colombia. Abbreviations: HCPIs: Healthcare Provider Institutions.

**Table 2.** Healthcare Fragmentation by Geographic Regions in Colombia according to the number of different providers involved during the first year of post-transplant healthcare attention.

| Geographic Region<sup>a</sup> | Mean | 95% CI |
| --- | --- | --- |
| Atlántica | 10.65 | 7.92 ; 13.38 |
| Bogotá DC | 8.25 | 5.51 ; 11.00 |
| Central | 10.79 | 7.90 ; 13.67 |
| Oriental | 9.07 | 6.42 ; 11.72 |
| Pacífica | 7.97 | 5.66 ; 10.28 |
| Other departments | 9.22 | 6.73 ; 11.72 |

<sup>a</sup> Atlantica: Atlántico, Bolívar, Cesar, Cordoba, Magdalena, La Guajira, Sucre, and San Andres; Metropolitan: Bogotá D.C. and Cundinamarca; Central: Antioquia, Caldas, Huila; Risaralda, Tolima, and Quindío; Oriental: Boyaca, Caquetá, Santander, Norte de Santander, Arauca, Meta, Casanare, and Guaviare; and Pacífica: Valle del Cauca, Cauca, Choco, and Nariño. Abbreviations: CI: Confidence Interval.

Patients were categorised into two groups: high fragmentation, comprising those who consulted 11 or more providers (≥p75), and low fragmentation, comprising those who consulted ten or fewer providers (<p75). 1,415 patients (69.7%) experienced low fragmentation, whereas 613 patients (30.2%) were exposed to high fragmentation. Table 3 shows the crude three-year mortality proportions for each baseline characteristic evaluated. Patients exposed to low fragmentation had a mortality proportion of 0.12, while those exposed to high fragmentation had a mortality proportion of 0.18 (p=0.04). Other characteristics where differences were observed included age category and CCI. No differences were found by sex or geographic region.

**Table 3.** Three-year Mortality Proportions by Baseline Characteristics.

| Characteristic | Mortality Proportion | 95% CI |
| --- | --- | --- |
| Fragmentation in the first year post-transplant |  |  |
| Low fragmentation (10 or fewer providers) | 0.12 | 0.10 - 0.14 |
| High fragmentation (11 or more providers) | 0.18 | 0.14 - 0.21 |
| Sex |  |  |
| Female | 0.14 | 0.12 - 0.16 |
| Male | 0.14 | 0.11 - 0.16 |
| Age category |  |  |
| 18-30 | 0.04 | 0.01 - 0.06 |
| 30-49 | 0.09 | 0.07 - 0.11 |
| 50-59 | 0.15 | 0.12 - 0.18 |
| 60-69 | 0.27 | 0.21 - 0.32 |
| 70 or older | 0.39 | 0.24 - 0.55 |
| Charlson Comorbidity Index |  |  |
| 3 or fewer | 0.10 | 0.08 - 0.11 |
| 4 to 5 | 0.23 | 0.19 - 0.28 |
| 6 to 7 | 0.25 | 0.17 - 0.33 |
| 8 or more | 0.14 | 0.05 - 0.23 |
| Geographic region |  |  |
| Atlantic | 0.09 | 0.05 - 0.14 |
| Bogotá DC | 0.16 | 0.13 - 0.19 |
| Central | 0.11 | 0.08 - 0.15 |
| Eastern | 0.16 | 0.10 - 0.22 |
| Pacific | 0.13 | 0.09 - 0.16 |
| Other departments | 0.00 | NA |
| Year of transplant |  |  |
| 2012 | 0.12 | 0.09 - 0.15 |
| 2013 | 0.11 | 0.08 - 0.14 |
| 2014 | 0.17 | 0.14 - 0.21 |
| 2015 | 0.15 | 0.10 - 0.19 |
| 2016 | 0.15 | 0.10 - 0.20 |

Abbreviations: 95% CI, 95% confidence interval.

Table 4 presents the multivariate Cox model, showing that exposure to a highly fragmented healthcare increases the risk of death at 3 years by 49% (HR: 1.49; 95% CI 1.12–1.97; p=0.01) after adjusting for sex, age, CCI, geographic region, year of transplantation, and insurer (Figure 3). Other factors associated with lower three-year survival included being over 30 years old, having a CCI greater than 4, and being female.

**Table 4.** Adjusted HR of overall survival of adults with kidney transplants by the level of healthcare fragmentation, defined by the number of different providers involved during the first year of post-transplant healthcare attention.

| Characteristic | Hazard Ratio | 95% CI | P-Value |
| --- | --- | --- | --- |
| Fragmentation in the first year post-transplant |  |  |  |
| Low fragmentation (10 or fewer providers) | Ref |  |  |
| High fragmentation (11 or more providers) | 1.49 | 1.12 - 1.97 | 0.01 |
| Sex |  |  |  |
| Male | Ref |  |  |
| Female | 1.30 | 1.01 - 1.67 | 0.04 |
| Age category |  |  |  |
| 18-30 | Ref |  |  |
| 30-49 | 2.30 | 1.15 - 4.60 | 0.02 |
| 50-59 | 3.88 | 1.93 - 7.80 | <0.01 |
| 60-69 | 6.84 | 3.40 - 13.77 | <0.01 |
| 70 or older | 12.01 | 5.51 - 26.20 | <0.01 |
| Charlson Comorbidity Index |  |  |  |
| 3 or fewer | Ref |  |  |
| 4 to 5 | 1.93 | 1.47 - 2.54 | <0.01 |
| 6 to 7 | 1.83 | 1.24 - 2.69 | <0.01 |
| 8 or more | 1.09 | 0.57 - 2.10 | 0.79 |
| Geographic region |  |  |  |
| Atlantic | Ref |  |  |
| Bogotá DC | 1.26 | 0.69 - 2.29 | 0.45 |
| Central | 0.93 | 0.51 - 1.71 | 0.82 |
| Eastern | 1.63 | 0.86 - 3.06 | 0.13 |
| Pacific | 1.12 | 0.61 - 2.08 | 0.71 |
| Other departments | 0.00 | 0.00 - . | 1.00 |
| Year of transplant |  |  |  |
| 2012 | Ref |  |  |
| 2013 | 0.95 | 0.65 - 1.39 | 0.78 |
| 2014 | 1.19 | 0.83 - 1.71 | 0.35 |
| 2015 | 1.18 | 0.78 - 1.78 | 0.43 |
| 2016 | 1.47 | 0.92 - 2.35 | 0.11 |

The HR was adjusted for Sex, Age, CCI, geographic region, year of transplant and insurer.
Abbreviations: 95% CI, 95% confidence interval; Ref: Reference.

**Fig 3.**
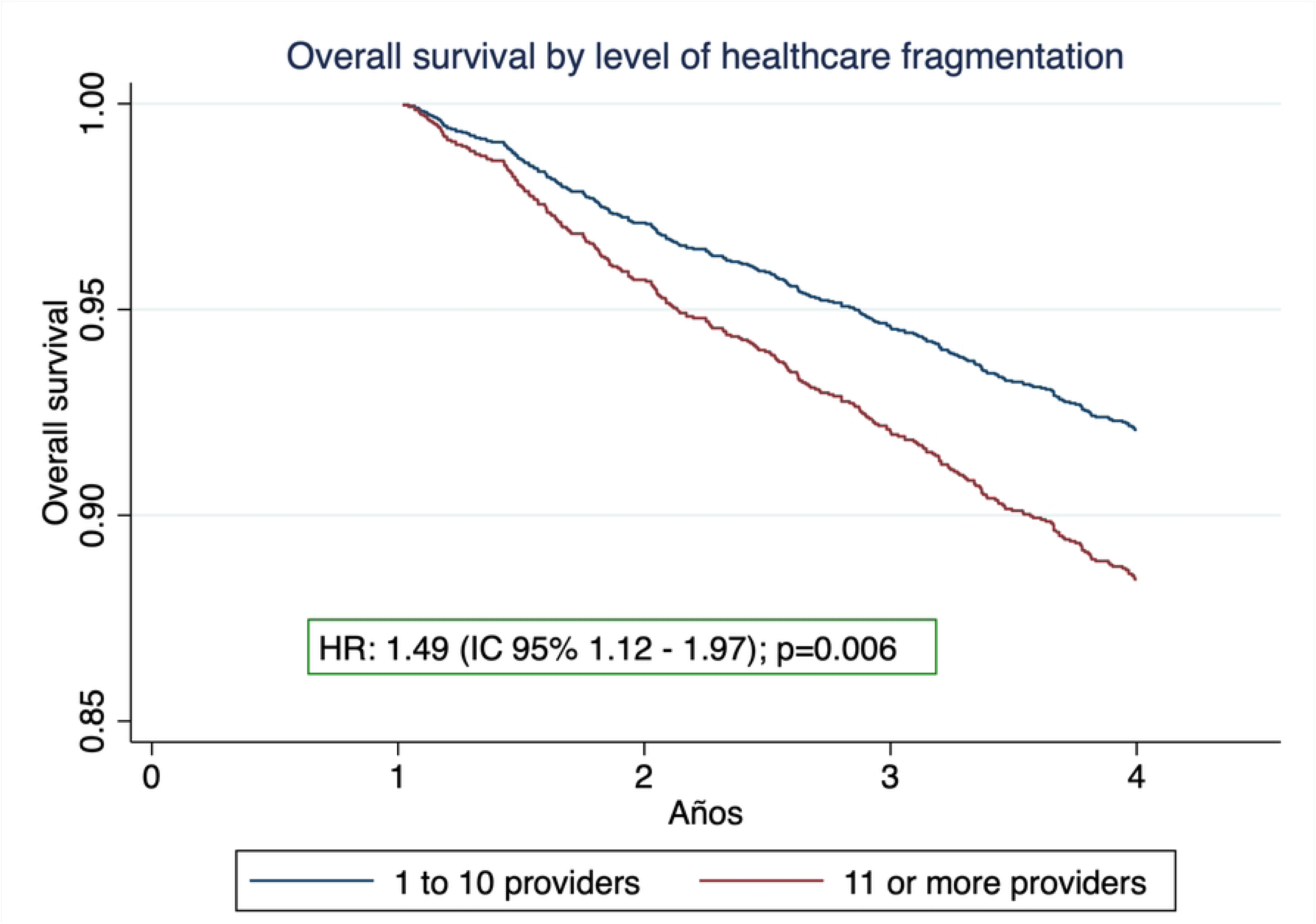
Three-year overall survival of adults with kidney transplants by the level of healthcare fragmentation, defined by the number of different providers involved during the first year of post-transplant healthcare attention.

## DISCUSSION

This study evaluated the association between healthcare fragmentation during the first year after kidney transplantation and survival outcomes. Our findings demonstrate that high fragmentation, defined as exposure to 11 or more unique healthcare providers within the first-year post-transplant, was associated with a 49% increased mortality risk within three years (HR: 1.49; 95% CI 1.12–1.97; p=0.01) for each additional provider participating in healthcare during the first-year post-transplant.

Healthcare fragmentation in this study was measured by the number of different healthcare institutions involved in a patient’s care during the first-year post-transplant. To our knowledge, this is the first study investigating the relationship between healthcare fragmentation and three-year survival of patients with kidney transplantation in an LMIC; however, this fragmentation metric has been used before in other chronic conditions, such as cancer, and our findings are consistent with the existing literature, also reporting poorer outcomes in patients with high levels of fragmentation [13–15,21]. Mechanisms driving these associations likely include delays in treatment, duplication of medical services, and poor coordination among healthcare providers, all of which undermine the potential benefits of transplantation [17].

Our results also revealed significant geographical variability in fragmentation across Colombia. Patients in the Central and Atlantic regions experienced the highest levels of fragmentation. In contrast, patients in the Pacific region exhibited lower fragmentation levels, likely due to limited access to specialised care rather than effective healthcare integration. Additionally, in areas with low population density and fewer healthcare facilities, patients often need to travel long distances to access specialised care, potentially leading to delays in treatment. These findings underscore the need for fragmentation metrics that account for travel distance and geographic barriers, particularly in countries with pronounced disparities in healthcare availability and accessibility.

This study has several strengths. The reliability of the data is a crucial advantage as it provides robust evidence on the relationship between healthcare fragmentation and transplant outcomes. The UPC database has been widely used in previous studies [23–30], and the RUAF death registry offers extensive coverage, documenting over 90% of deaths nationwide, further supporting their validity as valuable research tools. Additionally, the specificity of kidney transplant procedures, which are unlikely to be misclassified due to their cost and complexity, enhances the reliability of the data.

However, some limitations must be addressed. Administrative databases lack detailed clinical information, which may leave behind some crucial covariables. Furthermore, there needs to be a standardised measure of fragmentation that limits comparisons with studies from other parts of the world. Future research should explore more nuanced fragmentation metrics to capture its association with clinical outcomes better. Qualitative studies could also provide valuable insights into the experiences of patients and providers navigating fragmented healthcare systems, helping to inform policy and improve care integration.

## CONCLUSION

This study demonstrates that higher care fragmentation, measured by the number of unique healthcare providers involved during the first-year post-transplantation, is associated with reduced three-year overall survival in renal transplant patients in Colombia. Patients exposed to high fragmentation faced a 49% increased risk of death, highlighting the critical need to improve care coordination in this population. Our findings emphasise the importance of integrated healthcare models and multidisciplinary teams to reduce fragmentation, enhance clinical outcomes, and optimise resource use. Policymakers should prioritise efforts to streamline healthcare delivery, particularly in resource-limited settings, to mitigate the negative impacts of care fragmentation. Further research is needed to explore effective strategies for integrating care and addressing regional disparities in healthcare access, which could inform policies to improve outcomes for patients with chronic conditions, including those requiring renal transplantation.

## Data Availability

The following information sources: Single Registry of Enrollees, Mortality Registry Module from the Unified Affiliation Registry (RUAF) and Calculation Study of the Capitation Unit Database (Base del Estudio de Suficiencia de la Unidad Por Capitación, or UPC) are administered by the Colombian Ministry of Health and Social Protection. These databases are freely available upon request to the Technology of the Information and Communication Office of the Colombian Ministry of Health and Social Protection through the.

## ACKNOWLEDGMENTS

We thank Colombia’s Ministry of Health and Social Protection for providing the administrative databases that made this study possible. We also thank the Clinical Research Student Group at the Faculty of Medicine of the Universidad Nacional de Colombia for their valuable contributions. This study’s preliminary findings were presented and discussed within this group, enriching the interpretation and scope of our results. Additionally, this research was partially funded by the NIHR GHPSR researcher-led grant NIHR150067, which used UK aid from the UK Government to support global health research.

